# Increase in the Rate of Gut Carriage of Fluoroquinolone-Resistant *Escherichia coli* despite a Reduction in Antibiotic Prescriptions

**DOI:** 10.1101/2022.12.16.22283539

**Authors:** Veronika Tchesnokova, Lydia Larson, Irina Basova, Yulia Sledneva, Debarati Choudhury, Thalia Solyanik, Jennifer Heng, Teresa Christina Bonilla, Sophia Pham, Ellen M. Schartz, Lawrence T. Madziwa, Erika Holden, Scott J. Weissman, James D. Ralston, Evgeni V. Sokurenko

## Abstract

**Background:** Fluoroquinolone use for urinary tract infections has been steadily declining. Gut microbiota is the main reservoir for uropathogenic *Escherichia coli* but whether the carriage of fluoroquinolone-resistant *E. coli* has been changing is unknown.

**Methods:** We determined the frequency of isolation and other characteristics of *E. coli* nonsuceptible to fluoroquinolones (at ≥0.5 mg/L of ciprofloxacin) in 515 and 1605 *E. coli*-positive fecal samples collected in 2015 and 2021, respectively, from non-antibiotic-taking women of age 50+ receiving care in the Seattle area Kaiser Permanente Washington healthcare system.

**Results:** Between 2015 and 2021 the prescription of fluoroquinolones dropped nearly three-fold in the study population. During the same period, the rates of gut carriage of fluoroquinolone-resistant *E. coli* increased from 14.4 % to 19.9% (P=.005), driven by a significant increase of isolates from the recently emerged, pandemic multi-drug resistant clonal group ST1193 (1.7% to 4.3%; P=.007) and those with an incomplete set of or no fluoroquinolone-resistance determining mutations (2.3% to 7.5%; P<.001). While prevalence of the resistance-associated mobile genes among the isolates dropped from 64.1% to 32.6% (P<.001), co-resistance to third generation cephalosporins has increased 21.5% to 33.1%, P=.044).

**Conclusion:** Despite reduction in fluoroquinolone prescriptions, gut carriage of fluoroquinolone-resistant uropathogenic *E. coli* increased with a rise of previously sporadic lineages and co-resistance to third generation cephalosporins. Thus, to reduce the rates of antibiotic resistant urinary tract infections, greater focus should be on controlling the gut carriage of resistant bacteria.

**Short summary:** While prescription of fluoroquinolones dropped between 2015 and 2021, there was an increase in gut carriage of fluoroquinolone-resistant *Escherichia coli* among women of age 50+. Also, a rise of new resistant lineages and co-resistance to 3^rd^ generation cephalosporins occurred.

## Introduction

Antimicrobial resistance has reached pandemic proportions in the last few decades, increasing the mortality rates and health care costs associated with prescription of ineffective antibiotics [1, 2]. These developments prompted establishment of antimicrobial stewardship programs in health care institutions to minimize overuse of antibiotics and optimize appropriate antibiotic selection, dosing, and duration of therapy [3]. However, antibiotic-resistant bacteria are often circulating in the community as ‘commensal’ microbiota colonizing healthy individuals [4-6], and it remains unclear how antibiotic use reduction has affected their prevalence in that reservoir.

Urinary tract infections (UTIs) – bladder cystitis, pyelonephritis and urosepsis of both community and nosocomial origin – are among the most common reasons for antibiotic treatment [7]. Women of postmenopausal age -generally 50 years or older -are especially at high risk for severe and drug-resistant forms of UTI [5, 8]. UTIs are primarily caused by extra-intestinal pathogenic *Escherichia coli* that are mostly associated with strains from specific clonal groups [9, 10]. Resident gut bacteria carried by the patient as commensal microbiota are the primary reservoir of UTI-causing *E. coli*, including the drug resistant strains [5, 11-14].

Ciprofloxacin and other fluoroquinolones (FQ) had been for many years the most often prescribed antibiotics for UTI treatment and, commonly, for other infections [7, 15]. Against *E. coli* infections resistant to FQ and other antibiotics, third generation cephalosporins (3GC) are commonly used [16, 17]. Wide use of FQ has led to documentation of several concerning side effects including neurotoxicity and tendinitis as well as development of *Clostridium difficile* infections [18-20]. Most importantly, in the last two decades there has been a rampant rise in FQ resistant (FQR) pathogens in general and, specifically, FQR *E. coli* (FQREC) causing extra-intestinal infections [21]. The FQREC occurrence being strongly associated with the rates of hospitalization and mortality from sepsis [21]. The non-susceptibility to FQ is primarily associated with chromosomal point mutations in the so-called quinolone resistance-determining regions (QRDR) coding the main targets of FQ – bacterial DNA topoisomerases GyrA and ParC [22]. A set of at least three QRDR mutations – two in GyrA *and* one in ParC – typically results in the highest resistance level [23]. The FQR phenotype is also mediated by mobile, i.e transferrable, generally plasmid-mediated quinolone resistance (PMQR) genes [24, 25]. Mobile genes are also primarily responsible for resistance to 3GC, mostly mediated by extended-spectrum beta-lactamases (ESBL) [26].

To cutback the adverse effects associated with FQ and in attempt to reduce the spread of resistance, a campaign was launched in the mid-2010s to avoid prescription in patients with uncomplicated UTI and reserve FQ for more severe forms of UTIs [19, 27, 28]. However, it remains questionable whether a reduction in antibiotics use can be effective in reducing the rates of resistance in *E. coli* infections [29]. Moreover, theoretical models suggest that, once emerged, the resistant isolates can keep circulating as commensal colonizers even in the absence of antibiotics use [30, 31].

Here we examined the presence, clonal composition, FQR determinants and 3GC co-resistance of gut colonizing FQREC isolated from fecal specimens collected during 2015 and 2021 from non-antibiotic taking women of age 50+ enrolled into the Kaiser Permanente Washington (KPWA) health care system. While the study was not designed for a direct association analysis, we describe a significant increase in gut colonization by FQREC between 2015 and 2021 driven by a rise in specific bacterial lineages and 3GC co-resistance that occurred in parallel to the reduction in overall use of FQ but rise in the 3GC prescriptions among KPWA enrollees.

## Results

### Decline in the prescription of FQ and concurrent rise in the prescription of 3GC

Between 2010 and 2021, the number of enrollees in the Kaiser Permanente WA (KPWA) health plan (known as the GroupHealth Cooperative before 2018) of patients ≥18 yo ranged between ∼287K and ∼339K for total enrollment and ∼98K to ∼118K for women aged 50+ yo **(Figure 1A)**. Between 2010 and 2015, the fraction of total enrollees who were prescribed FQ remained relatively steady (6.40± 0.28%), but since 2016 the use of FQ declined >2.5-fold (to 2.30%, P < .001) by 2021 (**Figure 1B**).

**Figure 1.**
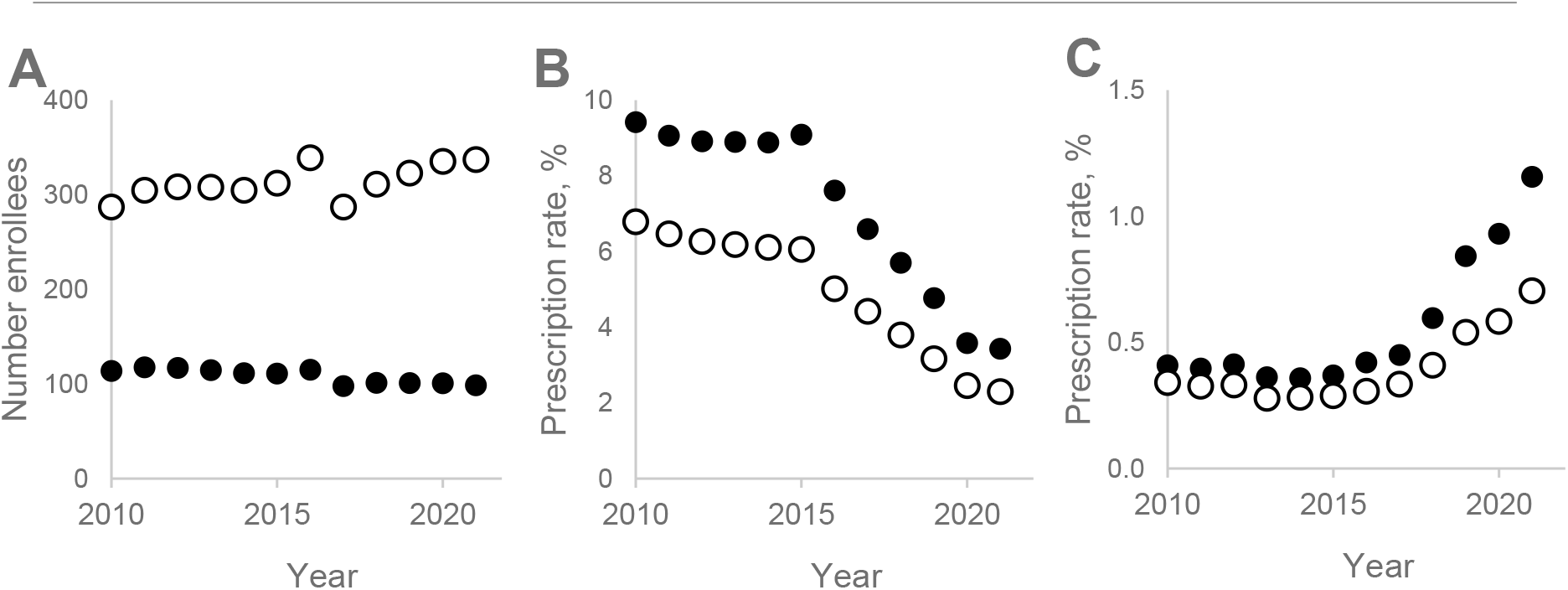
Antibiotics prescription among KPWA enrollees. (A) Total enrollment of 18+ yo in KPWA per year. (B) FQ yearly prescription rates. (C) 3CS yearly prescription. Open circles – all enrollees, closed circles – female enrollees which would be 50+ year old in 2021.

Among the target study population -women who reached age 50+ by 2021 – the use of FQ was significantly higher than in the total population in 2010-2015 (9.20± 0.39%, P < .001) but also declined as drastically (to 3.40%, P < .001) by 2021. The use of 3GC was proportionally much less than that of FQ but prescription rates increased from 0.30±0.04% in 2010-2015 to 0.70% by 2021 in the total population and from 0.40±0.05% to 1.13% in the target population (P < .001 for both) (**Figure 1C**). The temporal changes in the FQ and 3GC use reflected more general trends observed among all Medicare Part D enrollees across USA and, more prominently, in the Washington state (**Supplemental Figure S1**).

### The overall rate of gut carriage of FQREC increased due to a rise of certain isolate groups

Among fecal samples submitted in 2015 and 2021 from women of age 50+, the vast majority (89.6% and 90.3%, respectively; P=.623) yielded *E. coli*. Among the *E. coli*-positive samples from the 2015 study, 14.4% contained FQREC (**Table 1**). In the 2021 study, the rate of FQR isolates among the *E. coli* positive samples increased nearly 1.5-fold, to 19.9% (P=.005). In 95.9% and 95.0% of FQREC-positive samples in 2015 and 2021, respectively, resistant *E. coli* of only a single clonal group was identified per sample. The two most common FQR clonal groups remained the same in both studies – the pandemic multi-drug resistant ST131-*H30* and ST1193 (**Table 1**). While the combined carriage of the two pandemic clonal groups rose only from 7.8% in 2015 to 8.7% in 2021 (P=.098), carriage of ST1193 increased ∼2.5 fold (P=.007) and became equal to that of the previously dominant ST131-*H30*. All 2015 and 2021 isolates of ST1193 and, with one exception, ST131-*H30* had full set of at least three QRDR mutations – two in GyrA and one in ParC.

**Table 1.** Presence, clonality and QRDR mutations profile of fecal FQR *E. coli* isolated in 2015 and 2021.

| Category | 2015 |  | 2021 |  | P value <sup>b</sup> |
| --- | --- | --- | --- | --- | --- |
|  | No | % <sup>a</sup> | No | % <sup>a</sup> |  |
| **Total** | 575 |  | 1778 |  |  |
| ***E. coli* <sup>a</sup>** | 515 | 89.6 | 1605 | 90.3 | .623 |
| **FQR *E. coli* <sup>c</sup>** | 74 | 14.4 | 320 | 19.9 | **.005** |
| **ST131-H30** | 31 | 6.0 | 70 | 4.4 | .124 |
| **ST1193** | 9 | 1.7 | 69 | 4.3 | **.007** |
| **ST69 <sup>d</sup>** |  |  |  |  |  |
| **Total** | 7 | 1.4 | 51 | 3.2 | **.028** |
| **≥3 QRDR** | 4 | 0.8 | 8 | 0.5 | .464 |
| **2 QRDR** | 3 | 0.6 | 28 | 1.7 | **.040** |
| **1 QRDR** | 0 | 0.0 | 6 | 0.4 | .326 |
| **0 QRDR** | 0 | 0.0 | 9 | 0.6 | .087 |
| **Other <sup>d</sup>** |  |  |  |  |  |
| **Total** | 28 | 5.4 | 141 | 8.8 | **.012** |
| **≥3 QRDR** | 20 | 3.9 | 66 | 4.1 | .819 |
| **2 QRDR** | 1 | 0.2 | 5 | 0.3 | .662 |
| **1 QRDR** | 6 | 1.2 | 48 | 3.0 | **.022** |
| **0 QRDR** | 2 | 0.4 | 24 | 1.5 | **.047** |

<sup>a</sup> For '*E. coli*' category percent from all samples is given; for all other categories percent from only all *E. coli*-containing samples is calculated.
<sup>b</sup> Difference in prevalence between samples from 2015 and 2021 study was evaluated using two-tailed Fisher's exact test for 2x2 table; P values < .05 are in bold
<sup>c</sup> 3 samples in 2015 collection (1 sample with two clones - H30 and other ST, and two samples with four and two other ST clones), and 16 samples in 2021 collection (1 sample contained both ST131-H30 and ST1193, 2 samples contained ST131-H30 and ST69, 3 samples contained ST1193 and ST69, 1 sample contained ST1193 and other FQR clone, 3 samples contained ST69 and other FQR clone and 6 samples contained a mix of other FQR clones)
<sup>d</sup> Among other STs both 2015 and 2021 collections had ST131-H41, ST38, ST405, ST457, ST648, ST1163, as well as several STs from Clonal Complexes CC10, CC21, CC58; additionally, 2015 collection had isolates from ST156, ST410 and CC1196; 2021 collection had isolates belonging to ST95, CC14, ST636, ST349, ST394, ST354, etc. ST69 group represents a ST69 clonal complex.

The third most common FQREC in both studies were strains from one of the largest uropathogenic clonal groups, ST69, but its carriage rate increased >2 fold between 2015 and 2021 (P=.028) (**Table 1**). In contrast to ST131-*H30* and ST1193, only 15.7% of the 2021 ST69 isolates had ≥3 QRDR mutations and their overall increase was exclusively due to isolates with only two QRDR mutations (S83L of GyrA and S80I of ParC) (P=.040).

The remainder of FQREC belonged to various smaller, mostly uropathogenic clonal groups (a total of 21 in 2015 and 59 in 2021, Supplemental Table S2), with the combined carriage rate of such isolates increasing significantly in 2021 (P=.012). As with ST69, the increase of the FQREC from the smaller clonal groups was driven by the isolates with fewer than three QRDR mutations, but in this case was due to a significant rise of isolates with either a single mutation in GyrA (P=.022) or no mutation at all (P=.047) (**Table 1**).

### Isolates with FQ resistance-mediated mobile elements became less prevalent

Irrespectively of the QRDR mutations number, the majority of 2015 FQREC carried the PMQR genes (**Table 2**). In contrast, in 2021 isolates overall carriage of the mobile genes dropped ∼2-fold (P<.001). Though the PMQR elements remained (nearly) omnipresent among 2021 isolates with no QRDR mutations (97.2%), only a minority of the isolates with at least one QRDR mutation were carrying them. Especially notable drop was among the isolates with ≥3 QRDR mutations (23.7%; P<.001). In both years, most of isolates with the PMQR genes contained *qnrB* alone (Supplemental Table S3).

**Table 2.** Distribution of Plasmid-Mediated Quinolone Resistance (PMQR) determinants among FQR *E. coli* isolates with different numbers of QRDR mutations.

| No. QRDR mutations | Presence of PMQR determinants | 2015 |  | 2021 |  | P value |
| --- | --- | --- | --- | --- | --- | --- |
|  |  | No. isolates | % | No. isolates | % |  |
| no QRDR | YES | 2 | 100 | 35 | 97.2 | .811 |
|  | NO | 0 |  | 1 |  |  |
| 1 QRDR | YES | 4 | 66.7 | 16 | 29.1 | .063 |
|  | NO | 2 |  | 39 |  |  |
| 2 QRDR | YES | 2 | 50 | 8 | 24.2 | .273 |
|  | NO | 2 |  | 25 |  |  |
| ≥3 QRDR | YES | 42 | 63.6 | 51 | 23.9 | <.001 |
|  | NO | 24 |  | 162 |  |  |
| Total | YES | 50 | 64.1 | 110 | 32.6 | <.001 |
|  | NO | 28 |  | 227 |  |  |

### Increase in the prevalence of FQREC with a lower resistance level to FQ

We investigated how the presence of chromosomal QRDR mutations and PMQR-associated genes affected the ability of gut FQREC to grow at 0.5, 2.0 or 10.0 mg/L of the ciprofloxacin. The resistance level was directly correlated with the number of QRDR mutations, irrespective of clonal identity or the mobile gene presence. Nearly all isolates (277 out of 279; 99.3%) with at least three QRDR mutations were able to grow on plates with ciprofloxacin at a concentration of 10.0 mg/L (**Table 3**). At the same time, only 10 out of 136 isolates (7.4%; P<.001) having fewer than three QRDR mutations were able to grow at that concentration, with nearly all of them having 2 mutations. Majority of the remaining two-mutation isolates (85.7%) were able to grow at 2 mg/L, while only 26.2% (P<.001) of the isolates with a single QRDR mutation and 10.5% (P<.001) with no QRDR mutations were able to do so. In contrast to isolates with ≥3 as well as 2 QRDR mutations, most of isolates with 1 or, especially, no QRDR mutations (72.1% and 89.2%, respectively) were able to grow only at 0.5 mg/L of ciprofloxacin (P<.001). Due to the increase in prevalence of isolates with only 2 or fewer QRDR mutations in 2021, the overall proportion of FQREC isolates unable to grow at 10.0 mg/L increased from 17.7% in 2015 to 33.7% in 2021 (P=.006).

**Table 3.**
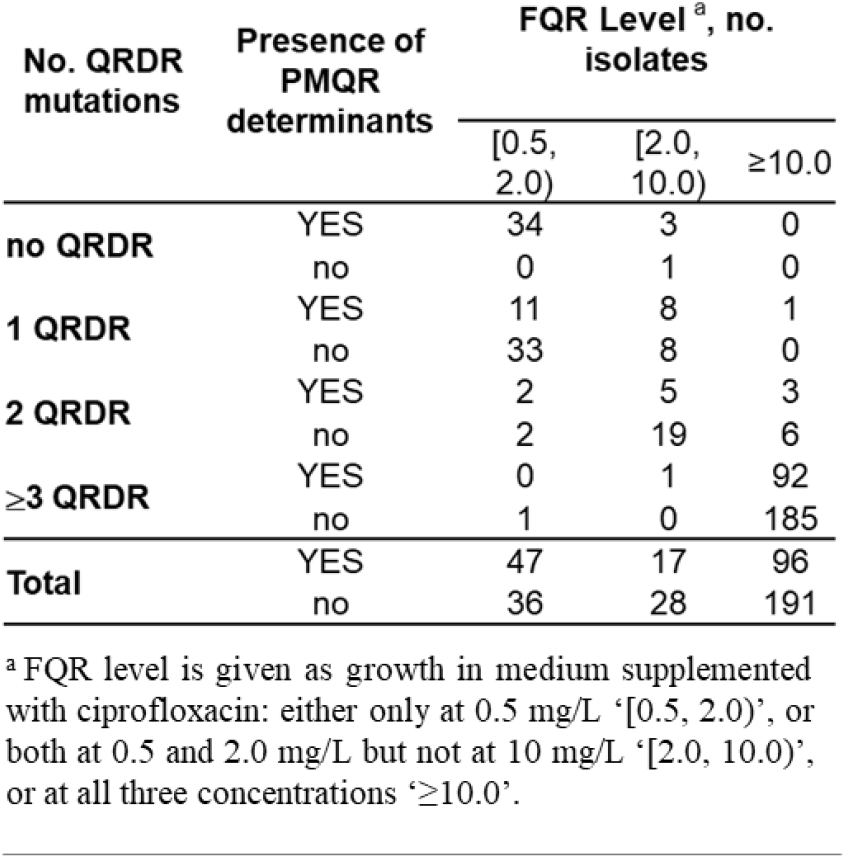
Relationship between level of resistance vs. number of QRDR mutations and presence of PMQR determinants in FQR *E. coli*.

| No. QRDR mutations | Presence of PMQR determinants | FQR Level<sup>a</sup>, no. isolates |  |  |
| --- | --- | --- | --- | --- |
| | | [0.5, 2.0) | [2.0, 10.0) | $\geq 10.0$ |
| no QRDR | YES | 34 | 3 | 0 |
|  | no | 0 | 1 | 0 |
| 1 QRDR | YES | 11 | 8 | 1 |
|  | no | 33 | 8 | 0 |
| 2 QRDR | YES | 2 | 5 | 3 |
|  | no | 2 | 19 | 6 |
| ≥3 QRDR | YES | 0 | 1 | 92 |
|  | no | 1 | 0 | 185 |
| Total | YES | 47 | 17 | 96 |
|  | no | 36 | 28 | 191 |

<sup>a</sup> FQR level is given as growth in medium supplemented with ciprofloxacin: either only at 0.5 mg/L '[0.5, 2.0)', or both at 0.5 and 2.0 mg/L but not at 10 mg/L '[2.0, 10.0)', or at all three concentrations ' $\geq 10.0$ '.

### Resistance to 3GC has increased among the gut FQREC

The gut carriage rate of *E. coli* co-resistant to FQ and 3GC rose >2-fold between 2015 and 2021 (3.1% vs 6.8%; P=.002), with their proportion among FQREC isolates rising from 21.5% to 33.1% (P=.044) **(Figure 2)**. 3GC resistance increased numerically across different categories of FQREC, but most significantly – from 18.5% to 48.1% (P=.006) -among isolates with at least three QRDR mutations. Among the 3GC co-resistant isolates from either 2015 or 2021, the majority were ESBL producers (94.1% vs. 75.0%, respectively; P=.079).

**Figure 2.**
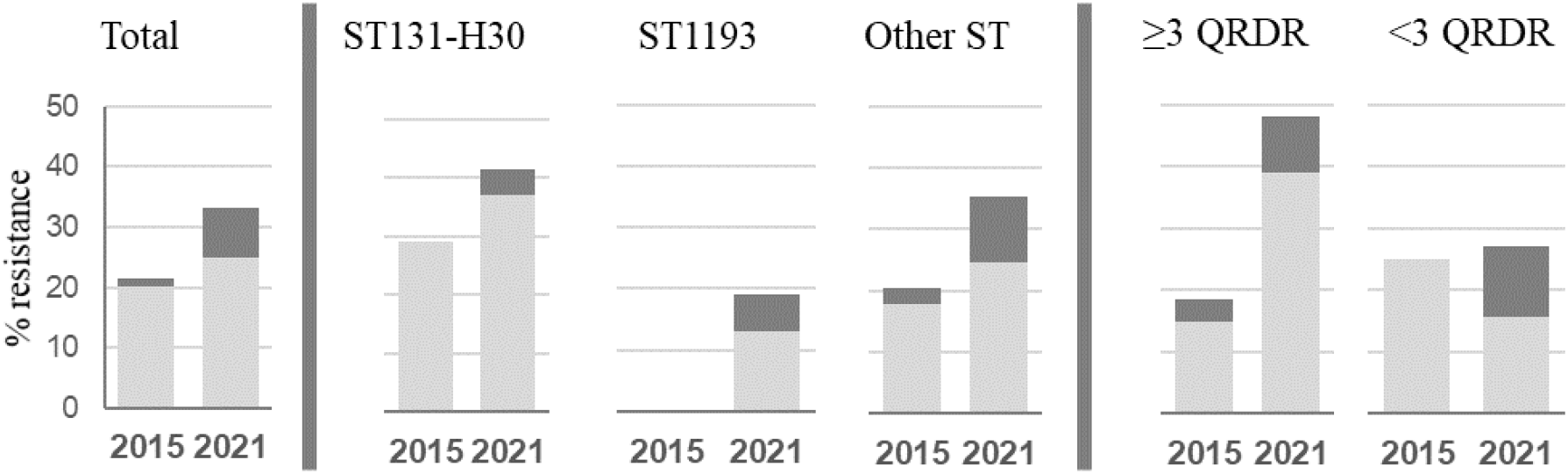
proportion of 3GC-resistant strains among FQREC. Distribution of total 3GC-resistance (darl grey) and ESBL (light) among FQR *E. coli* isolates from 2015 and 2021 *E. coli* collections, total, belonging to different clones (St131-*H30*, ST1193 and all other), or harbouring different number of QRDR mutations.

## Discussion

The commensal gut microbiota is the primary reservoir of UTI-causing *E. coli*, with gut carriage profiles mirroring UTI from the perspectives of clonal composition and drug-resistance profiles of *E. coli* as well as patient age demographics [32, 33]. In particular, elderly women are colonized more often by FQREC and are at a higher risk for FQR forms of UTI than younger women [5]. As recently as 2016, FQs were the most prescribed antibiotic for treating UTIs [34]. However, accumulated evidence for adverse effects combined with a sharp rise in FQ resistance of uropathogenic enterobacteria led to FDA, CDC and IDSA recommendations for curbing FQ use in the treatment of uncomplicated UTI [19, 27, 28, 35, 36]. Though, FQ continues to be recommended for patients who have no alternative treatment options, complicated UTIs, pyelonephritis or other severe infections where the benefits of FQ outweigh the risks of adverse effects [17, 20, 36]. Thus, resistance to FQ in uropathogens remains a significant clinical problem, and reduction in gut carriage of FQREC would be a welcome development. This is especially important because FQR phenotype of *E. coli* has the strongest association with UTI-caused bacteremia in individuals aged over 50 yo and overall sepsis mortality rates in adults [37, 38].

Here we show that the use of FQ was significantly higher among women of age 50+ compared to all enrollees in the KPWA healthcare system, potentially reflecting the higher incidence of UTIs in this category of women. In both populations, however, FQ prescription significantly declined between 2015 and 2021. While our study was not designed to establish a direct casual correlation between the changes in antibiotics use and resistance level, certain temporal changes in the profiles of gut FQREC might be potentially linked to the drop in FQ use. These include the higher prevalence of gut *E. coli* with a reduced (but still clinically relevant) resistance level (due to presence fewer QRDR mutations) and the overall reduction in isolates carrying the resistance-mediating mobile genes. Surprisingly, however, the total rate of gut carriage of fully resistant clonal groups with a complete set of the QRDR mutations remained at least the same, and the prevalence of FQREC from the pandemic multi-drug resistant clonal group ST1193 significantly increased. As a result, the rate of the overall gut carriage of FQR *E. coli* significantly increased in 50+ yo women between 2015 and 2021, despite the decline in FQ prescriptions. This and the fact that women in our study did not use any antibiotics for at least one year before providing the fecal sample suggests that community circulation of gut colonizing FQREC can be fully sustained even in the absence of antibiotic use, consistent with some predictions [30, 31].

For the last 20 years, clonal group ST131-*H30* has been globally predominant among FQREC and multi-drug resistant *E. coli* [8, 13, 39]. Starting a decade ago, however, another UTI-associated clonal group of FQREC - ST1193 - has been on the rise world-wide [40, 41]. We show here that the ST1193 gut carriage rate went up between 2015 and 2021 and is now on par with ST131-*H30*. It is worrisome because ST1193 appears to be more pathogenic than ST131-*H30* as it more frequently causes UTIs in younger women with a relatively robust host defense system [11, 42]. It is unclear whether the higher virulence or other fitness attributes are linked to the significant rise in gut carriage, but if ST1193 continues to increase in the coming years, the result could be an increase of infections caused by it even if the tendency to reduce prescriptions of FQ continues. Similarly, another FQREC clonal group that is on the rise in gut carriage, ST69, is known to more frequently cause UTI in children to whom it could be transmitted [43, 44].

Bacterial topoisomerases GyrA and ParC are the main targets of FQ, and acquisition of QRDR mutations is the major mechanism of *E. coli* becoming non-susceptible to FQ [45]. It is provocative to suggest that reduced usage of FQ could create a selective environment allowing FQREC strains with just few QRDR mutations to circulate in greater numbers. However, the rise in prevalence of FQREC from ST69 was due to isolates that carry two out of three QRDR mutations – one in GyrA and one in ParC. This combination of mutations has been rarely observed in previous studies and it was proposed that, upon the acquisition of the first mutation in GyrA, there is a weak resistance gain by a mutation in ParC without the simultaneous occurrence of a second mutation in GyrA [22, 46-48]. Importantly, a simultaneous double mutation is a very low-probability genetic event, which should be a limiting factor for emergence of the fully resistant FQREC during the treatment with high FQ doses [23]. Thus, the rise of FQREC with two QRDR mutations could present a significant clinical problem, because the two-mutation strains are only one mutation away from becoming highly resistant. This simple genetic event could be easily selected in a patient during antibiotic therapy, resulting in treatment failure and infection relapse. Further studies are needed, however, to determine the clinical significance of the two-mutation strains and genetic backgrounds underlying their rise among the ST69 strains.

The prevalence of isolates carrying mobile PMQR genes was reduced between 2015 and 2021, suggesting that the selective pressure needed to maintain the plasmids became weaker. However, this reduction was noted only among isolates with QRDR mutations, while carriage of the mobile resistance genes remained essential for the FQREC isolates without the mutations. While the impact of the mobile resistance elements is less pronounced quantitatively than that of the QRDR mutations, considering the rise of FQREC without QRDR mutations between 2015 and 2021, the role of mobile elements in the FQ non-susceptibility remains clinically significant.

Finally, there was an increase in gut carriage of FQREC strains that are also resistant to 3GC. The primary but not exclusive mechanism of 3GC resistance *in E. coli* is production of ESBL, which is a diverse class of enzymes dominated by CTX-M family typically coded by plasmid-associated genes [49]. In our analysis, 3GC resistance increased across all FQREC categories and clonal groups, suggesting that this had happened under a generally imposed selective pressure. Especially troublesome is the doubling of 3GC resistance among the isolates with ≥3 QRDR mutations, i.e., highly resistant to FQ. Notably, the prescription of 3GC was significantly increased in KPWA enrollees between 2015-2021, especially among women 50+ yo. The reason for the increase is not fully understood but may potentially be associated with 3GC being used more often as a replacement choice for FQ. Though a direct correlation between the rise in the 3GC co-resistance and increased use of the antibiotic was not investigated here, it is plausible to suggest that these are interconnected developments.

Taken together, results of our study suggest that, while increased use of antibiotics in patients can lead to emergence of resistant isolates, the latter can continue to spread in the community even if antibiotic use is decreased. In turn, the continuous level of community circulation of the resistant bacteria could lead to a sustained rate of antimicrobial therapy-refractory infections, consistent with a recent study [29]. Therefore, on one hand our study fully supports the efforts of antimicrobial stewardship to decrease overuse of antibiotics. On the other hand, reduction in specific antibiotic prescriptions may not alone be a sufficient measure to reduce the spread of resistance. Another factor to consider is the use of FQ in animals like poultry farming and its spread into the environment that was not accounted for in our study but might have a significant effect on the circulation of resistant bacteria in the community [50]. One way or another, our study suggests that besides the reduction of antibiotic use, identification and/or selective de-colonization of carriers may provide an effective way to control resistance in clinical infections. The decolonization measures could be especially effective if implemented in high-risk patients or community groups and/or against the most clinically dangerous resistant strains. This might be achieved, for example, via the use of probiotic strains, bacteriophages or both [51-54]. However, a better understanding of both epidemiology and ecology of antibiotic resistant bacteria is needed to identify the basis of their fitness as colonizers and the best way to target them. Thus, we propose that there may be a need to expand the role of antimicrobial stewardship programs from being focused only on antibiotic use in clinical settings to being also oriented towards screening for and decolonization of commensal carriage of antibiotic resistant strains in the most vulnerable individuals.

## Methods

### Study design and participants

This study on collection and analysis of fecal and urine samples from women without documented prescribed antibiotics was approved by the Kaiser Permanente Washington (KPWA) Research Institute Institutional Review Board and was originally carried out between July 2015 and January 2016, described here [5], and then repeated under the same protocol between May and December 2021.

### Processing of fecal samples and typing of fecal E. coli

Samples were evaluated visually for the quality of fecal matter, plated on three UTI agar plates (plain or supplemented with ciprofloxacin at 0.5, 2 or 10 mg/L, then split into two tubes (with and without 10% glycerol) and stored at -80°C. Plates were incubated 16-20h at 37°C, and up to 30 single colonies (SCs) morphologically identified as potential *E. coli* were cultured, saved, tested for ESBL production, and typed. ESBL production was determined by growth on plates with cefoxitin 4 and 8 mg/L (CEF-4 and CEF-8), ceftazidime 8 and 16 mg/L (CAZ-8 and CAZ-16) and on HardyCHROM ESBL HDx (Hardy Diagnostics, USA). *E. coli* clonality was determined by CH typing based on *fumC*/*fimH* sequencing [55], presence of QRDR mutations was determined by sequencing of *gyrA* and *parC* [13]. All reactions were carried out by 2-step PCR where additional nested PCR is used to get highly specific single bands with T7-tails primers. TMQR determinants were identified in multiplex PCR as described previously [56]. The multiplex is intended to identify 9 TMQR determinants: *qnrA, qnrD, qnrB, qnrS, oqxAB, aac(6′)-Ib-cr, qepA, qnrC* and *qnrVC*. All primers are listed in Supplemental Table S1.

### Prescription data collection

All KPWA enrollees (enrolled at least 10 months within a year) were selected for years 2010-2021 and their EMRs were checked for prescription of FQ or 3GC class antibiotic at least once during the year. The number of female enrollees who would have been 50+ yo in 2021 were identified and their prescription rates were calculated separately.

### Statistical analysis

Prevalence of FQREC and individual clones among 2015 and 2021 isolates was compared by Fisher’s exact test. Trends in FQ and 3GC prescription rate was analyzed using linear regression. Analysis was carried out using STATA 14.2 Software (StataCorp, Texas, USA).

## Data Availability

All data produced in the present work are either contained in the manuscript or are available upon request to the authors.

## Acknowledgements

We would like to thank Drs. Steve Moseley, Paul Thottingal, Jessoca Zering, Katarina Kameran, Marisa D’Angeli and Yuan-Po Tu for the critical reading of manuscript and suggestions for additional data to improve it, and Arina Pushkina and Denys Davydenko for excellent technical assistance.

## Supplemental Materials

**Table S1.**
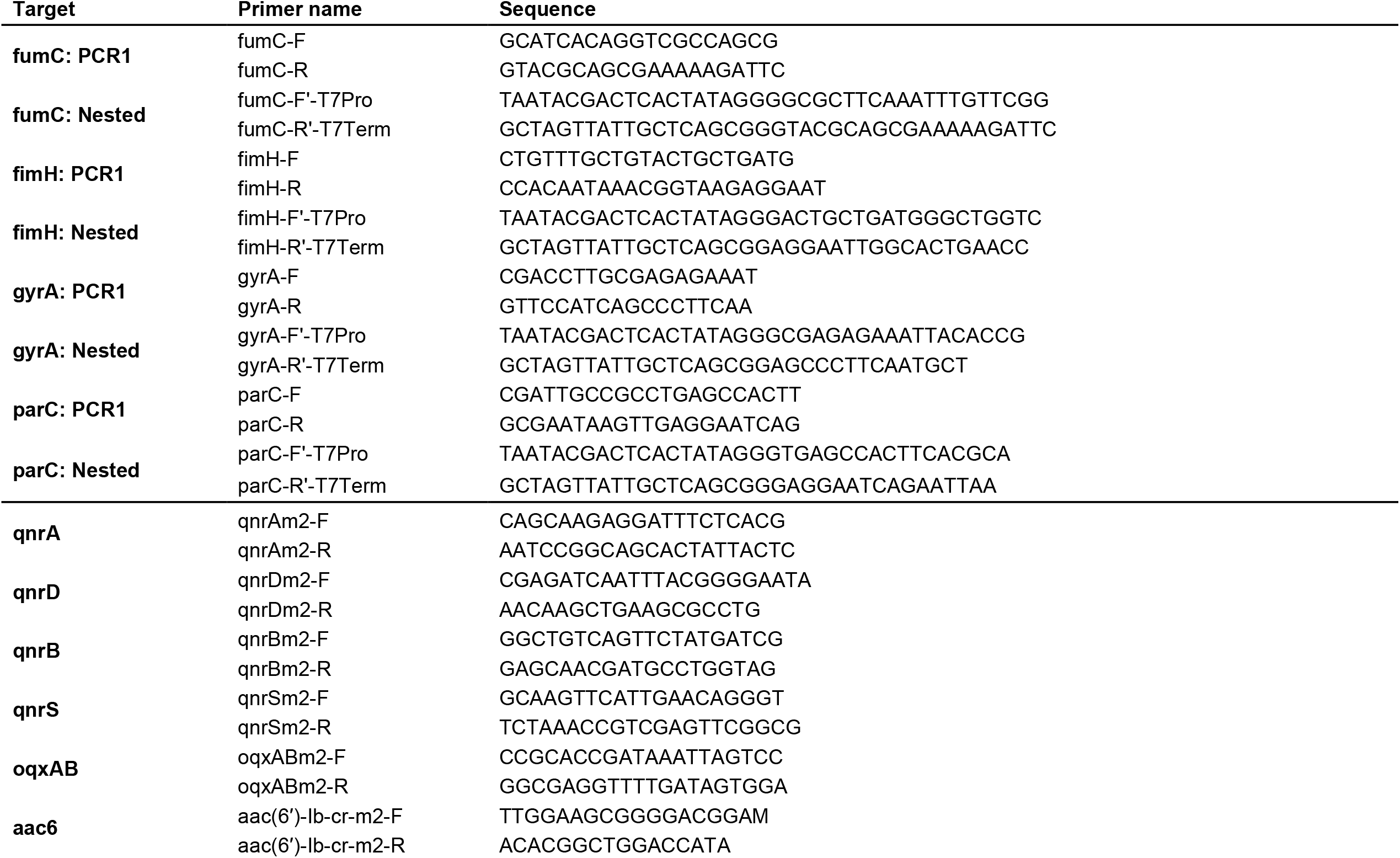
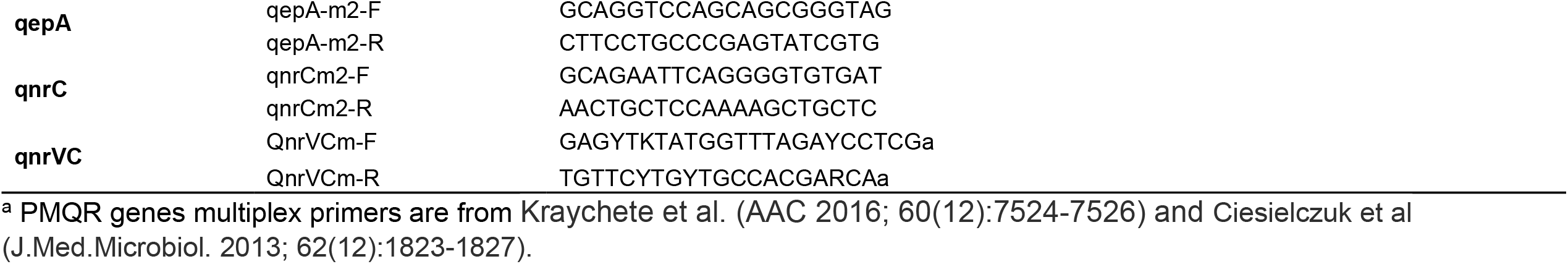
Primers used for CH typing, gyrA-parC sequencing and PMQR genes multiplex ^a^

**Table S2.**
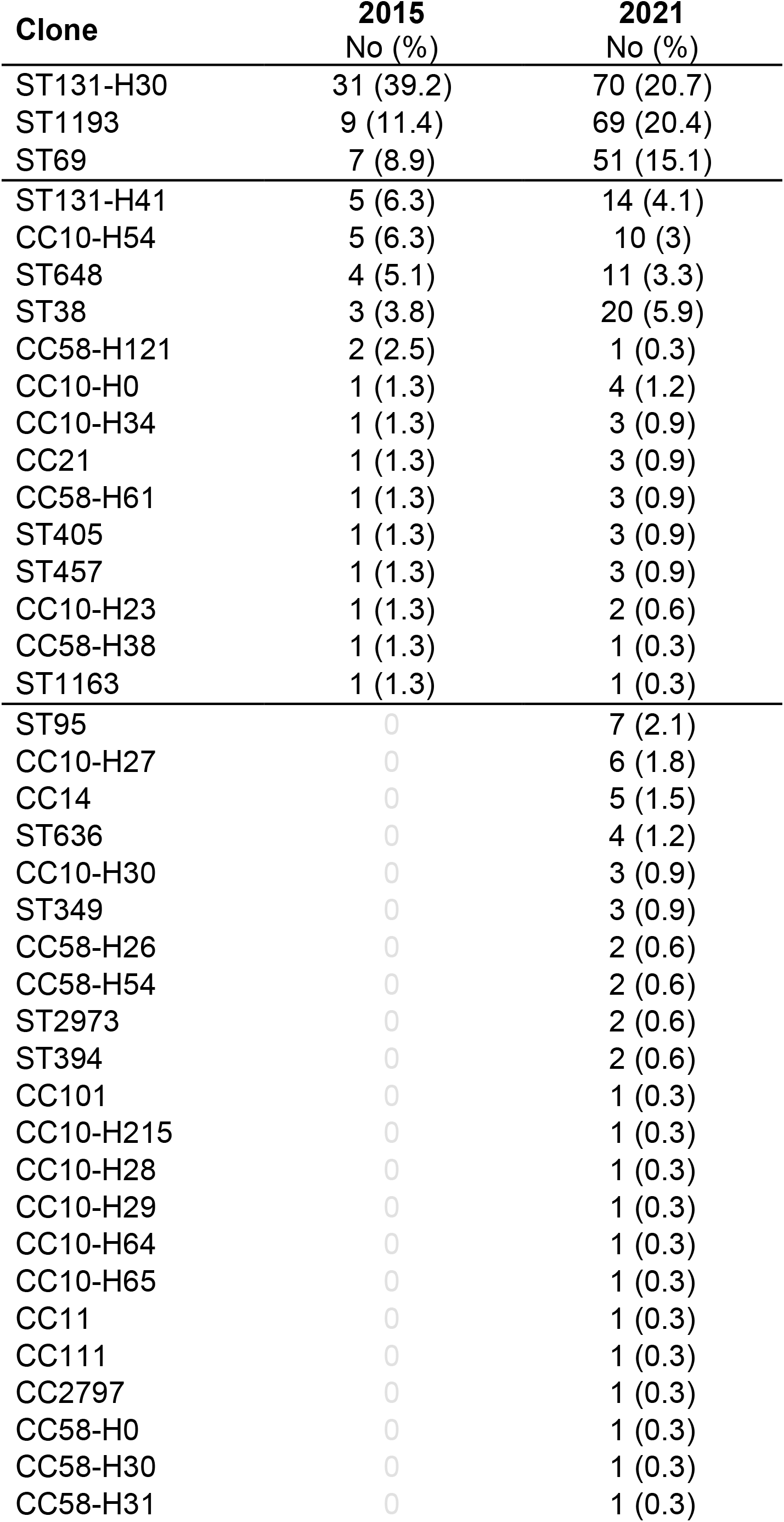
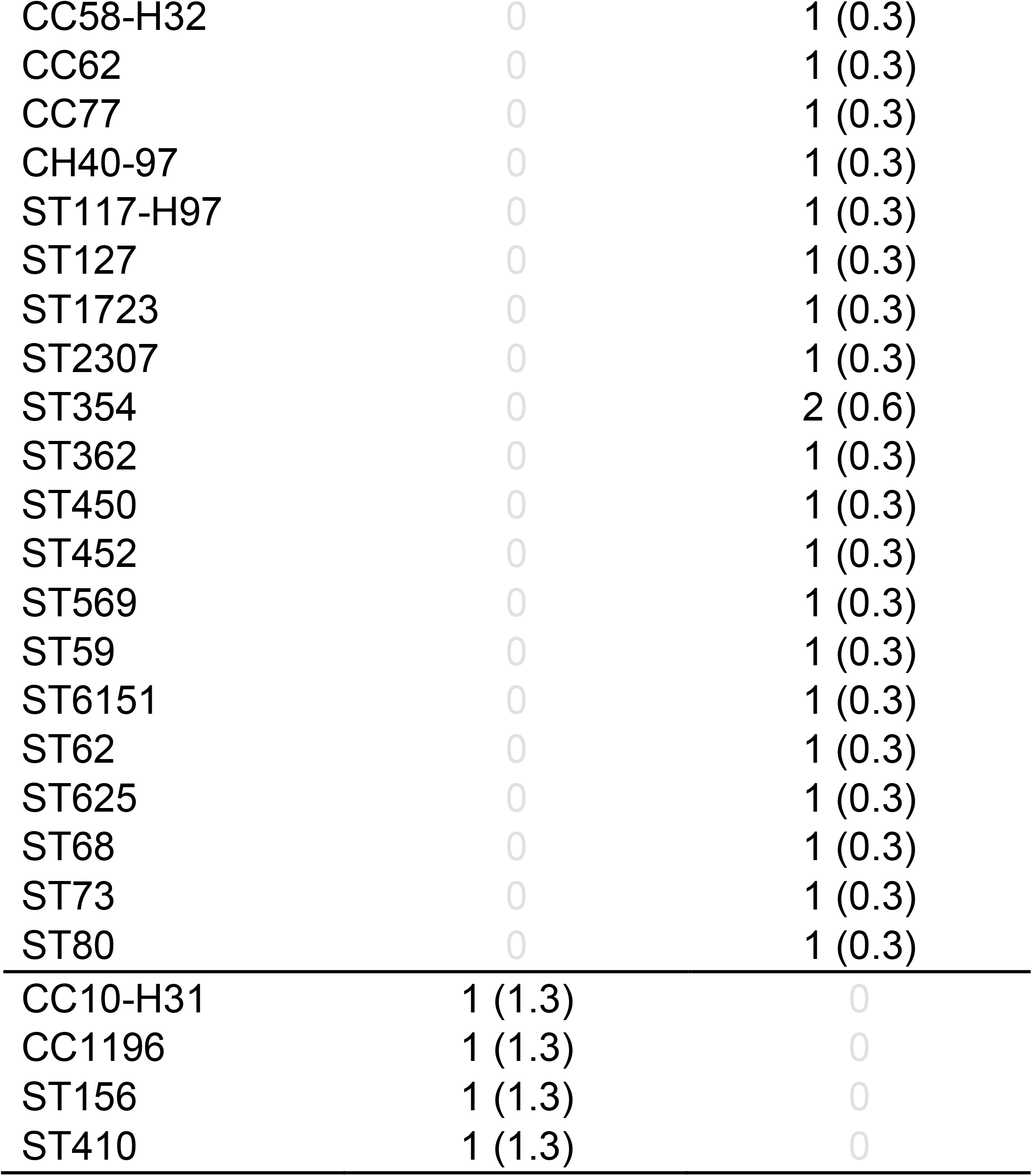
FQR *E. coli* clones identified in 2015 and 2021 studies.

**Table S3.** PMQR determinants identified in 2015 and 2021 FQR *E. coli* isolates ^a^

| **PMQR type** | **2015** | **2021** |
| --- | --- | --- |
| qnrB + qepA | 2 | 2 |
| qnrB | 30 | 62 |
| aac6 | 4 | 14 |
| qnrS | 7 | 20 |
| qnrD | 1 | 0 |
| qnrB + qnrS | 1 | 4 |
| qepA + aac6 | 1 | 0 |
| qnrB + oqxAB | 1 | 0 |
| qnrB + qepA + oqxAB | 2 | 0 |
| qepA | 0 | 4 |
| qnrS + oqxAB | 0 | 2 |
| qnrB + aac6 | 0 | 2 |
| qnrB + qepA + aac6 | 1 | 0 |
| Total | 50 | 110 |

<sup>a</sup> aac6 stands for aac(6')-Ib-cr

**Supplementary Figure S1.**
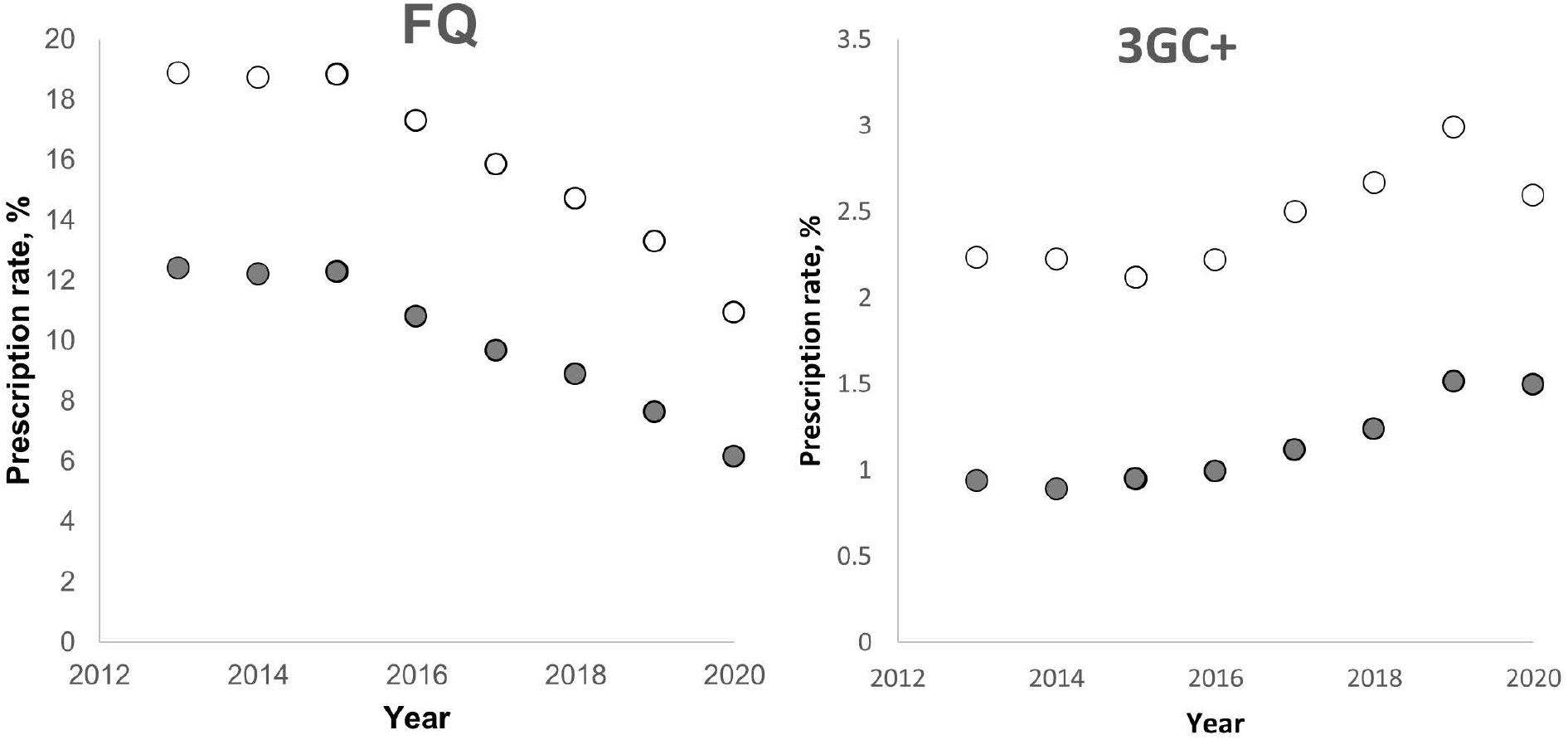
Prescription rates of fluoroquinolones (FQ) and third and higher generation cephalosporins (including carbapenems) (3GC+) among Medicare enrollees in 2013-2020 in Washington state (closed circles) and USA (open circles).

The rate is calculated as percent of total beneficiaries (the total number of unique Medicare Part D beneficiaries with at least one claim for the drug per year) from total number of enrollees per year. All data were downloaded from http://data.cms.gov:

- The CMS Program Statistics - Medicare Part D tables provide use and Part D drug costs by type of Part D plan (stand-alone prescription drug plan and Medicare
- The CMS Program Statistics - Medicare Part D Enrollment tables provide data on characteristics of the Medicare Part D covered population.

